# Clinico-Epidemiological Characteristics, Hospital Care Interventions, and Clinical Outcomes of Patients with SARS-CoV-2 in Nepal: The Mediating Role of Age

**DOI:** 10.64898/2026.09.08.26362589

**Authors:** Mamata Sharma Neupane, Hafizah Che Hassan, Surendra Uranw, Raj Kumar Mehta

## Abstract

Coronavirus disease 2019 (COVID-19) outcomes among hospitalized patients are influenced by demographic characteristics, clinical presentation, and requirements for hospital-based care. Age is an important determinant of COVID-19 outcomes; however, its statistical role in the relationships between hospital-care interventions and survival has received limited attention in resource-constrained settings. This prospective observational analytical study assessed clinico-epidemiological characteristics, hospital-care interventions, clinical outcomes, and the mediating role of patient age group among adults hospitalized with SARS-CoV-2 infection in Nepal. A total of 348 adults with laboratory-confirmed SARS-CoV-2 infection were consecutively recruited at a tertiary care teaching hospital from March 30, 2022 to June 30, 2023 and followed until discharge or in-hospital death. Associations with survival were examined using Chi-square/Fisher’s exact tests and multivariable binary logistic regression. Mediation analyses were performed using Hayes’ PROCESS Macro version 4.2, Model 4, with 5,000 bias-corrected bootstrap samples. Of 348 participants, 277 (79.6%) survived and 71 (20.4%) died. Older adults constituted 41.1% of participants. Hypertension (29.3%) and diabetes mellitus (23.0%) were the most common comorbidities, while fever (81.0%), cough (75.9%), and dyspnea (48.0%) were the predominant symptoms. Age group and dyspnea remained independently associated with hospital survival in multivariable analysis. Steroid therapy, oxygen therapy, and respiratory support were significantly associated with survival in unadjusted analyses. Age group partially statistically mediated the steroid therapy–survival relationship (indirect effect = −0.1371; 95% bootstrap CI: −0.3053 to −0.0154), while the direct effect remained significant (c′ = −2.8670; p = 0.0049). No significant age-mediated indirect effects were observed for oxygen therapy or respiratory support. These findings highlight the importance of age and respiratory presentation in hospital outcomes and suggest age-related heterogeneity in the steroid therapy–survival relationship. The mediation finding should be interpreted as statistical rather than causal because age precedes treatment exposure.

## 1. INTRODUCTION

Coronavirus disease 2019 (COVID-19), caused by severe acute respiratory syndrome coronavirus 2 (SARS-CoV-2), created an unprecedented global public health emergency, placing sustained pressure on health systems, hospital capacity, critical care services, and healthcare resources worldwide [1]. Although the acute global emergency phase has ended, SARS-CoV-2 continues to circulate and cause hospitalization and mortality, particularly among older and clinically vulnerable populations [1]. The pandemic also exposed longstanding inequalities in preparedness and access to essential hospital resources, including medical oxygen, respiratory support, critical care services, and evidence-based therapeutics. These challenges were particularly important in low- and middle-income countries (LMICs), where resource constraints could influence both the delivery of hospital care and patient outcomes [2].

The clinical course of COVID-19 is heterogeneous, ranging from mild or asymptomatic infection to severe pneumonia, hypoxemic respiratory failure, multiorgan dysfunction, and death. Among hospitalized patients, outcomes are influenced by an interplay of demographic characteristics, pre-existing comorbidities, presenting clinical manifestations, disease severity, and the level of supportive and therapeutic care required during hospitalization [3,4]. Large observational studies have consistently demonstrated that advancing age is among the strongest predictors of severe COVID-19 and mortality. For example, the OpenSAFELY study involving more than 17 million adults demonstrated a pronounced age gradient in COVID-19-related mortality even after adjustment for multiple demographic and clinical characteristics [3]. Age therefore represents more than a descriptive characteristic of hospitalized populations; it is an important factor in risk stratification, clinical decision-making, resource utilization, and interpretation of treatment– outcome relationships.

Hospital management of moderate, severe, and critical COVID-19 evolved considerably during the pandemic. Supplemental oxygen, systemic corticosteroids, and different forms of respiratory support became central components of care for patients with hypoxemia and progressive respiratory failure [5,6]. The RECOVERY trial established that dexamethasone reduced 28-day mortality among hospitalized patients receiving oxygen or invasive mechanical ventilation, while no mortality benefit was demonstrated among patients who did not require respiratory support [5]. These findings subsequently informed international recommendations supporting systemic corticosteroids for patients with severe or critical COVID-19 [6]. Oxygen therapy and escalation of respiratory support similarly remained essential components of clinical management for patients developing respiratory compromise.

However, interpreting associations between hospital-care interventions and survival in observational studies is challenging. Treatments such as corticosteroids, oxygen therapy, and respiratory support are not administered randomly in routine clinical practice; they are more likely to be provided to patients with greater disease severity or higher clinical risk. Consequently, an intervention may appear to be associated with an adverse outcome because patients receiving it were already more severely ill—a problem commonly described as confounding by indication. Patient characteristics that influence both clinical management and prognosis may further complicate these relationships. Age is particularly relevant because older adults are more vulnerable to severe disease and adverse outcomes and may differ from younger patients in their need for hospital-based interventions, physiological reserve, burden of comorbidity, and response to acute illness [3,7]. Understanding the relationship between treatment and outcome therefore requires consideration not only of *whether* an intervention is associated with survival but also of the pathways through which such associations may operate.

This issue is particularly important in LMIC health systems. The COVID-19 pandemic demonstrated substantial global inequalities in access to medical oxygen, respiratory support, intensive care capacity, trained personnel, and other resources required for the management of severe respiratory illness [2]. Evidence generated predominantly in high-income settings may therefore not fully represent patient profiles, patterns of care, resource availability, or outcomes in resource-constrained health systems. Context-specific hospital-based evidence is essential for understanding how demographic and clinical characteristics interact with the delivery of care and survival in these settings.

Nepal experienced multiple waves of COVID-19 while simultaneously facing limitations in hospital beds, critical care capacity, medical oxygen supply, equipment, and specialized human resources. Available studies from Nepal have described the clinical characteristics and mortality patterns of patients with COVID-19 and have indicated that older age, comorbidities, and respiratory manifestations are prominent among patients with adverse outcomes [8,9]. A multicentre assessment of COVID-19-related deaths across hospitals in Nepal, for example, reported a substantial burden of mortality among older patients and those with underlying comorbidities [8]. However, much of the available Nepalese evidence has been descriptive or has focused on identifying factors associated with mortality. Comparatively less attention has been directed toward understanding how patient characteristics may influence the relationship between hospital-care interventions and survival.

Conventional association and multivariable regression analyses are valuable for identifying factors independently associated with clinical outcomes, but they primarily estimate adjusted relationships between predictors and outcomes. They do not, by themselves, establish whether an observed relationship operates through an intermediate statistical pathway. Mediation analysis extends conventional regression by decomposing an exposure–outcome relationship into direct and indirect effects through a specified mediator [10,11]. This approach can therefore provide additional insight into the statistical pathways underlying observed relationships. In the context of COVID-19 hospital care, mediation analysis offers an opportunity to investigate whether the association between a hospital-care intervention and survival differs through an age-related pathway rather than treating age solely as a conventional covariate.

The role of age in such a pathway warrants particular investigation. Age has been extensively studied as a predictor, risk factor, stratification variable, and potential confounder in COVID-19 outcome research [3,4]. Far less evidence has examined age within a formal mediation framework linking hospital-care interventions with survival, particularly in LMIC settings. Importantly, because chronological age precedes both hospitalization and treatment, its inclusion as a mediator in observational data should be interpreted as **statistical mediation rather than proof of a causal biological mediation mechanism**. Such analysis can nevertheless be useful for decomposing observed treatment–outcome associations and examining whether these relationships differ through pathways represented by patient age [10,11]. This distinction is important for avoiding causal overinterpretation while allowing a more detailed assessment of treatment–outcome relationships than conventional association analyses alone.

There is therefore a need for context-specific evidence that integrates the clinico-epidemiological characteristics of hospitalized patients, hospital-care interventions, survival outcomes, and pathway-based statistical analysis within a resource-constrained healthcare setting. Such evidence may improve understanding of the heterogeneity of hospitalized COVID-19 populations, strengthen age-sensitive risk stratification, and inform interpretation of hospital-care outcomes in Nepal and comparable LMIC settings.

Accordingly, this study aimed to assess the demographic and clinico-epidemiological characteristics, hospital-care interventions, and clinical outcomes of adults hospitalized with SARS-CoV-2 infection at a tertiary care hospital in Nepal; identify demographic and clinical factors associated with hospital survival; and examine whether patient age statistically mediated the relationships between three principal hospital-care interventions—steroid therapy, oxygen therapy, and respiratory support—and survival outcome at hospital discharge. By integrating conventional outcome analysis with mediation analysis, the study sought to move beyond description of treatment–outcome associations and provide additional insight into age-related pathways underlying hospital survival among patients with COVID-19 in a resource-constrained setting.

## 2. MATERIALS AND METHODS

### Ethics statement

Ethical approval for the study was obtained from the Research Ethics Committee of Lincoln University College, Malaysia (LUC/INT/113/2022; 7 February 2022), the Nepal Health Research Council, Kathmandu, Nepal (NHRC/77/2022 PhD; 23 March 2022), and the Institutional Review Committee of Chitwan Medical College Teaching Hospital, Nepal (CMC-IRC/078/079-149; 30 March 2022).

The subsequent amendment incorporating the mediation objective and analysis using Hayes’ PROCESS Macro was reviewed and approved by the Research Ethics Committee of Lincoln University College (LUC/REC/INT/211/2023; 17 August 2023). The amendment did not alter participant recruitment, clinical management, data collection procedures, or participant risk.

Written informed consent was obtained from participants before enrolment. When patients lacked decision-making capacity because of critical illness, unconsciousness, mechanical ventilation, or another clinical condition, written informed consent was obtained from their Legally Authorized Representative. Participation was voluntary, and refusal to participate did not affect clinical care. Participant information was handled confidentially, and only de-identified data were used for statistical analysis.

### Study design and setting

A prospective observational analytical study was conducted among adult patients hospitalized with laboratory-confirmed SARS-CoV-2 infection at Chitwan Medical College Teaching Hospital (CMCTH), Bharatpur, Nepal, from March 30, 2022 to June 30, 2023. Participants were prospectively followed from hospital admission until a definitive hospital outcome of discharge alive or in-hospital death.

CMCTH is a tertiary-level teaching and referral hospital providing specialized medical, surgical, diagnostic, emergency, and critical care services. During the COVID-19 pandemic, the hospital operated designated COVID-19 wards, isolation facilities, and intensive care units for the management of patients with varying levels of disease severity. These facilities provided hospital-based management including corticosteroid therapy, supplemental oxygen, and different levels of respiratory support according to patients’ clinical requirements and institutional treatment protocols.

The study was observational; no treatment or other aspect of routine clinical management was assigned or modified by the investigators. The reporting of this study was guided by the Strengthening the Reporting of Observational Studies in Epidemiology (STROBE) recommendations.

### Study population and eligibility criteria

The source population consisted of adult patients admitted to the COVID Ward, COVID Intensive Care Unit, or Isolation Ward of CMCTH during the study period. Patients were eligible if they were aged ≥18 years, had SARS-CoV-2 infection confirmed by reverse transcription polymerase chain reaction (RT-PCR), were admitted primarily for management of COVID-19, and provided written informed consent. For patients who were unconscious, critically ill, mechanically ventilated, or otherwise unable to provide consent at enrolment, written informed consent was obtained from a Legally Authorized Representative.

Patients aged <18 years, patients with suspected but unconfirmed SARS-CoV-2 infection, patients admitted primarily for conditions other than COVID-19, and patients with incomplete essential clinical information required for the principal study analyses were excluded. Patients transferred to another healthcare institution before a definitive hospital outcome could be established were also excluded.

Each participant was followed throughout hospitalization until either discharge following clinical recovery or in-hospital death. No post-discharge or long-term follow-up was undertaken.

### Sample size and sampling

The required sample size was determined for the study’s multivariable analytical objective using the logistic regression sample-size approach described by Hsieh and colleagues [12,13]. The calculation assumed a significance level of 5%, statistical power of 80%, an expected event proportion of 20%, and an anticipated odds ratio of 0.70 for a covariate one standard deviation above the mean. Based on these assumptions, the required sample size was 348 participants.

A non-probability consecutive sampling technique was used. All eligible hospitalized patients meeting the predefined criteria were approached consecutively during the study period until the required sample size was achieved. Consecutive recruitment was selected to reduce arbitrary participant selection and to capture the spectrum of hospitalized SARS-CoV-2 cases presenting to the study hospital during the study period.

A total of 348 participants were enrolled and followed to a definitive hospital outcome. Complete follow-up was achieved for all participants, with no loss to follow-up, withdrawals, or missing outcome data.

### Study variables and operational definitions

The primary dependent variable was **hospital survival outcome**, defined as the participant’s status at the end of hospitalization and categorized as survived/discharged alive or died during hospitalization.

The demographic and background characteristics evaluated included age, sex, marital status, province of residence, smoking habit, and alcohol consumption. Clinico-epidemiological variables included pre-existing comorbidities, presenting clinical symptoms, and clinical complications documented during hospitalization.

The principal **hospital-care interventions** were steroid therapy, oxygen therapy, and respiratory support. These variables were determined from physician orders, medication administration records, nursing documentation, oxygen therapy records, and respiratory support records and were categorized according to whether the participant received the respective intervention during hospitalization. These interventions were provided as part of routine clinical care according to clinical indications and were not assigned by the investigators.

For the mediation analyses, steroid therapy, oxygen therapy, and respiratory support were separately specified as independent variables (X); **patient age group** was specified as the mediator (M); and hospital survival outcome was specified as the dependent variable (Y).

Chronological age was obtained from the hospital admission record and recorded in completed years. Age was subsequently categorized into the predefined young-adult, middle-adult, and old-adult groups used consistently throughout the study. Because chronological age is a fixed characteristic that precedes hospital treatment, age group was treated as an **exploratory statistical mediator**, and the resulting indirect effects were not interpreted as demonstrating a causal mechanism whereby hospital treatment altered patient age.

### Data collection instrument and procedures

Data were collected prospectively using a structured case record form developed for the study. The instrument was designed to obtain sociodemographic characteristics, clinico-epidemiological information, hospital-care and treatment information, and clinical outcome at hospital discharge. Information was obtained from participant interviews where applicable and from hospital source documents, including admission records, physician progress notes, nursing records, medication administration charts, oxygen therapy records, respiratory support documentation, and discharge or death records.

The case record form was reviewed for content and face validity and was pretested among a small group of hospitalized patients with confirmed SARS-CoV-2 infection who met the study eligibility criteria but were not included in the final analytical sample. Pretesting assessed the clarity and completeness of the instrument, feasibility of data extraction, operational definitions, coding procedures, and time required for data collection. Minor modifications were made before commencement of the main study.

Eligible patients were identified following hospital admission. After confirming eligibility and obtaining informed consent, baseline demographic and clinical information was recorded. Participants were subsequently monitored throughout their hospital stay, and treatment-related information and clinical events were documented prospectively. Final survival status was recorded at hospital discharge or death.

### Data quality assurance

Research assistants involved in data collection received standardized training from the principal investigator before commencement of the study. Training included study objectives, eligibility criteria, participant recruitment, informed consent procedures, consent from Legally Authorized Representatives, operational definitions, completion of the case record form, extraction of information from hospital records, confidentiality, and standardized data collection procedures.

The principal investigator supervised data collection on a daily basis. Completed forms were checked for completeness and consistency, and information was verified against original hospital records where necessary. Discrepancies were resolved through re-examination of the relevant source documents and, when appropriate, clarification with the treating clinical team.

Before analysis, data were checked for coding errors, out-of-range values, inconsistencies, and completeness. Range and logical consistency checks were performed, and identified discrepancies were resolved by referring to the original case record forms and hospital records. Complete hospital outcome information was available for all 348 participants; therefore, no imputation of missing outcome data was required.

### Statistical analysis

Data were coded, entered, cleaned, and analyzed using **IBM SPSS Statistics version 25**. Categorical variables were summarized using frequencies and percentages, while continuous variables, where applicable, were summarized using appropriate measures of central tendency and dispersion.

Bivariate analyses were performed to examine unadjusted associations between demographic, clinico-epidemiological, hospital-care variables, and hospital survival outcome. Pearson’s Chi-square test was used for categorical variables when its assumptions were satisfied. Fisher’s Exact test was used when expected cell frequencies were <5.

Multivariable binary logistic regression was performed to identify demographic and clinical factors independently associated with hospital survival while controlling for potential confounding. Variables demonstrating statistical significance in bivariate analyses together with variables considered clinically relevant were entered simultaneously into the multivariable model. Associations were expressed as adjusted odds ratios (AORs) with corresponding 95% confidence intervals (CIs) and p-values.

Laboratory and molecular variables and their associated regression models were **not included in the analyses reported in the present manuscript**.

### Mediation analysis

Mediation analysis constituted a principal analytical component of the study. The statistical mediating role of patient age group in the association between hospital-care interventions and hospital survival was examined using **Hayes’ PROCESS Macro version 4.2, Model 4**.

Three separate mediation models were evaluated:

1. Steroid therapy (X) → age group (M) → hospital survival (Y);
2. Oxygen therapy (X) → age group (M) → hospital survival (Y); and
3. Respiratory support (X) → age group (M) → hospital survival (Y).

For each model, PROCESS was used to estimate the total effect (c), direct effect after inclusion of the mediator (c′), and indirect effect through age group (a × b). Indirect effects were evaluated using **5,000 bias-corrected bootstrap resamples**, thereby avoiding reliance on the assumption of a normally distributed indirect effect. Statistical evidence of mediation was considered present when the 95% bootstrap confidence interval for the indirect effect did not include zero.

Because age necessarily precedes the hospital-care interventions in temporal sequence, the mediation models were interpreted as **exploratory statistical decomposition of the observed intervention–outcome associations rather than causal mediation models**. Thus, an indirect effect through age should not be interpreted as indicating that hospital treatment caused a change in patient age. Rather, the analysis was intended to quantify the extent to which age statistically accounted for variation in the observed relationship between each hospital-care intervention and survival.

All statistical tests were two-tailed, and a p-value <0.05 was considered statistically significant.

## 3. RESULTS

### Participant characteristics and clinical outcomes

A total of 348 adults with laboratory-confirmed SARS-CoV-2 infection were consecutively enrolled and included in the final analysis. All participants were followed from hospital admission until discharge alive or in-hospital death, with no loss to follow-up and complete outcome data.

Older adults constituted the largest age group (143/348, 41.1%), followed by middle-aged adults (119/348, 34.2%) and young adults (86/348, 24.7%). More than half of the participants were male (188/348, 54.0%), and 192 (55.2%) were residents of Bagmati Province.

Hypertension was the most prevalent pre-existing comorbidity, affecting 102 (29.3%) participants, followed by diabetes mellitus in 80 (23.0%) and chronic obstructive pulmonary disease (COPD) in 44 (12.6%). Thyroid disorders, chronic kidney disease, and chronic liver disease were less common. Fever (282/348, 81.0%), cough (264/348, 75.9%), and dyspnea (167/348, 48.0%) were the predominant presenting symptoms (Table 1).

**Table 1.**
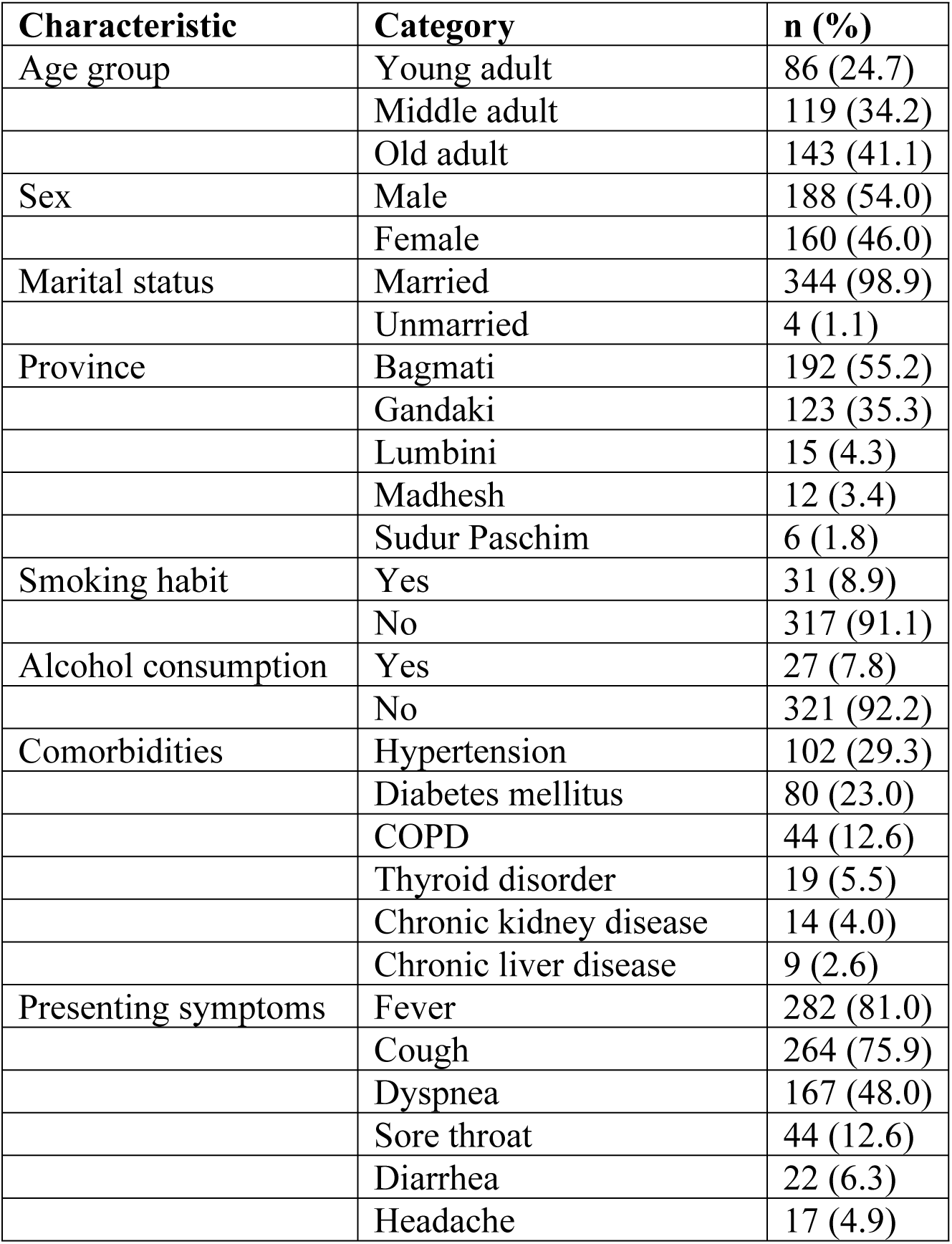
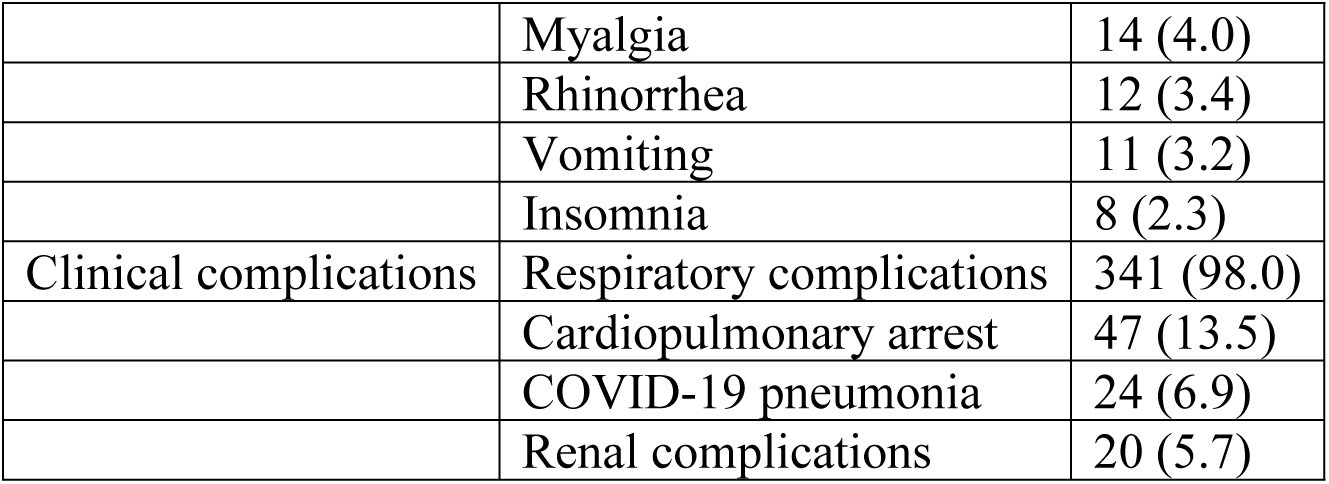
Sociodemographic and clinico-epidemiological characteristics of hospitalized patients with SARS-CoV-2 infection (N = 348)

| Characteristic | Category | n (%) |
| --- | --- | --- |
| Age group | Young adult | 86 (24.7) |
|  | Middle adult | 119 (34.2) |
|  | Old adult | 143 (41.1) |
| Sex | Male | 188 (54.0) |
|  | Female | 160 (46.0) |
| Marital status | Married | 344 (98.9) |
|  | Unmarried | 4 (1.1) |
| Province | Bagmati | 192 (55.2) |
|  | Gandaki | 123 (35.3) |
|  | Lumbini | 15 (4.3) |
|  | Madhesh | 12 (3.4) |
|  | Sudur Paschim | 6 (1.8) |
| Smoking habit | Yes | 31 (8.9) |
|  | No | 317 (91.1) |
| Alcohol consumption | Yes | 27 (7.8) |
|  | No | 321 (92.2) |
| Comorbidities | Hypertension | 102 (29.3) |
|  | Diabetes mellitus | 80 (23.0) |
|  | COPD | 44 (12.6) |
|  | Thyroid disorder | 19 (5.5) |
|  | Chronic kidney disease | 14 (4.0) |
|  | Chronic liver disease | 9 (2.6) |
| Presenting symptoms | Fever | 282 (81.0) |
|  | Cough | 264 (75.9) |
|  | Dyspnea | 167 (48.0) |
|  | Sore throat | 44 (12.6) |
|  | Diarrhea | 22 (6.3) |
|  | Headache | 17 (4.9) |
|  | Myalgia | 14 (4.0) |
|  | Rhinorrhea | 12 (3.4) |
|  | Vomiting | 11 (3.2) |
|  | Insomnia | 8 (2.3) |
| Clinical complications | Respiratory complications | 341 (98.0) |
|  | Cardiopulmonary arrest | 47 (13.5) |
|  | COVID-19 pneumonia | 24 (6.9) |
|  | Renal complications | 20 (5.7) |

Abbreviation: COPD, chronic obstructive pulmonary disease. Comorbidities, symptoms, and complications were not mutually exclusive.

Regarding hospital care, 82.2% of participants received steroid therapy, 224 (64.4%) received supplemental oxygen, and 138 (39.7%) required respiratory support. All participants received antibiotic therapy. Overall, 277 (79.6%) participants survived to hospital discharge and 71 (20.4%) died during hospitalization (Table 2).

**Table 2.** Hospital-care interventions and clinical outcomes among hospitalized patients with SARS-CoV-2 infection (N = 348)

| Variable | Category | n (%) |
| --- | --- | --- |
| Steroid therapy | Yes | 286 (82.2) * |
|  | No | 62 (17.8) |
| Antibiotic therapy | Yes | 348 (100.0) |
|  | No | 0 (0.0) |
| Oxygen therapy | Yes | 224 (64.4) |
|  | No | 124 (35.6) |
| Respiratory support | Yes | 138 (39.7) |
|  | No | 210 (60.3) |
| Hospital outcome | Survived | 277 (79.6) |
|  | Died | 71 (20.4) |

### Factors associated with hospital survival

Survival differed significantly across age groups (p < 0.001). Mortality was 4.7% among young adults, compared with 30.3% among middle-aged adults and 21.7% among older adults. Sex was also associated with survival (p = 0.021), with mortality occurring in 25.0% of males compared with 15.0% of females.

Among pre-existing comorbidities, COPD was significantly associated with survival outcome (p = 0.002). Mortality occurred in 4.5% of participants with documented COPD compared with 22.7% among those without COPD. Hypertension, diabetes mellitus, chronic liver disease, chronic kidney disease, and thyroid disorders were not significantly associated with survival in the unadjusted analyses.

Among presenting symptoms, dyspnea demonstrated a strong association with outcome (p < 0.001). Of the participants presenting with dyspnea, 49/167 (29.3%) died compared with 22/181 (12.2%) among those without dyspnea. Myalgia and headache were also statistically associated with survival in unadjusted analyses; however, these findings involved small cell frequencies and should be interpreted cautiously.

COVID-19 pneumonia and cardiopulmonary arrest were strongly associated with mortality (both p < 0.001). All 24 participants documented as having COVID-19 pneumonia died, while all 47 participants experiencing cardiopulmonary arrest died (Table 3). Because these events occurred during the clinical course and showed complete separation of outcomes, they were considered indicators of severe disease progression rather than appropriate baseline predictors for multivariable prognostic modelling.

**Table 3.** Unadjusted associations of sociodemographic and clinico-epidemiological characteristics with hospital survival among patients with SARS-CoV-2 infection (N = 348)

| Characteristic | Category | Survived, n (%) | Died, n (%) | $\chi^2$ value | p-value |
| --- | --- | --- | --- | --- | --- |
| Age group | Young adult | 82 (95.3) | 4 (4.7) | 20.391 | <0.001† |
|  | Middle adult | 83 (69.7) | 36 (30.3) |  |  |
|  | Old adult | 112 (78.3) | 31 (21.7) |  |  |
| Sex | Male | 141 (75.0) | 47 (25.0) | 5.323 | 0.021* |
|  | Female | 136 (85.0) | 24 (15.0) |  |  |
| Smoking habit | Yes | 31 (100.0) | 0 (0.0) | 8.723 | <0.001† |
|  | No | 246 (77.6) | 71 (22.4) |  |  |
| Alcohol consumption | Yes | 27 (100.0) | 0 (0.0) | 7.503 | 0.002†* |
|  | No | 250 (77.9) | 71 (22.1) |  |  |
| Hypertension | Yes | 81 (79.4) | 21 (20.6) | 0.003 | 0.956 |
|  | No | 196 (79.7) | 50 (20.3) |  |  |
| Diabetes mellitus | Yes | 64 (80.0) | 16 (20.0) | 0.010 | 0.919 |
|  | No | 213 (79.5) | 55 (20.5) |  |  |
| COPD | Yes | 42 (95.5) | 2 (4.5) | 7.799 | 0.002†* |
|  | No | 235 (77.3) | 69 (22.7) |  |  |
| Chronic liver disease | Yes | 8 (88.9) | 1 (11.1) | 0.491 | 0.421† |
|  | No | 269 (79.4) | 70 (20.6) |  |  |
| Chronic kidney disease | Yes | 11 (78.6) | 3 (21.4) | 0.009 | 0.570† |
|  | No | 266 (79.6) | 68 (20.4) |  |  |
| Thyroid disorder | Yes | 12 (63.2) | 7 (36.8) | 3.345 | 0.068 |
|  | No | 265 (80.5) | 64 (19.5) |  |  |
| Fever | Yes | 222 (78.7) | 60 (21.3) | 0.700 | 0.403 |
|  | No | 55 (83.3) | 11 (16.7) |  |  |
| Cough | Yes | 210 (79.5) | 54 (20.5) | 0.002 | 0.966 |
|  | No | 67 (79.8) | 17 (20.2) |  |  |
| Sore throat | Yes | 38 (86.4) | 6 (13.6) | 1.420 | 0.233 |
|  | No | 239 (78.6) | 65 (21.4) |  |  |
| Dyspnea | Yes | 118 (70.7) | 49 (29.3) | 15.799 | <0.001* |
|  | No | 159 (87.8) | 22 (12.2) |  |  |
| Myalgia | Yes | 14 (100.0) | 0 (0.0) | 3.739 | 0.038†* |
|  | No | 263 (78.7) | 71 (21.3) |  |  |
| Diarrhea | Yes | 20 (90.9) | 2 (9.1) | 1.850 | 0.136† |
|  | No | 257 (78.8) | 69 (21.2) |  |  |
| Vomiting | Yes | 11 (100.0) | 0 (0.0) | 2.912 | 0.078† |
|  | No | 266 (78.9) | 71 (21.1) |  |  |
| Rhinorrhea | Yes | 12 (100.0) | 0 (0.0) | 3.186 | 0.062† |
|  | No | 265 (78.9) | 71 (21.1) |  |  |
| Headache | Yes | 17 (100.0) | 0 (0.0) | 4.581 | 0.019†* |
|  | No | 260 (78.5) | 71 (21.5) |  |  |
| Insomnia | Yes | 8 (100.0) | 0 (0.0) | 2.099 | 0.158† |
|  | No | 269 (79.1) | 71 (20.9) |  |  |

Footnote: $\chi^2$ = Chi-square test; COPD = chronic obstructive pulmonary disease; †Fisher's exact test applied where appropriate; \*statistically significant at $p < 0.05$ .

### Hospital-care interventions and survival

Steroid therapy, oxygen therapy, and respiratory support were each significantly associated with survival in unadjusted analyses (all p < 0.001). Mortality was higher among patients who received these interventions than among those who did not. In particular, 71/224 (31.7%) patients receiving supplemental oxygen died, whereas no deaths occurred among the 124 patients who did not require oxygen therapy (Table 4).

**Table 4.** Unadjusted association of hospital-care interventions with hospital survival among patients with SARS-CoV-2 infection (N = 348)

| Hospital-care intervention | Category | Survived n (%) | Died n (%) | $\chi^2$ | p-value |
| --- | --- | --- | --- | --- | --- |
| Steroid therapy | Yes | 196 (73.7) | 70 (26.3) | 16.400 | <0.001† |
|  | No | 61 (98.4) | 1 (1.6) |  |  |
| Oxygen therapy | Yes | 153 (68.3) | 71 (31.7) | 49.378 | <0.001† |
|  | No | 124 (100.0) | 0 (0.0) |  |  |
| Respiratory support | Yes | 67 (48.6) | 71 (51.4) | 135.737 | <0.001† |
|  | No | 210 (100.0) | 0 (0.0) |  |  |

† Fisher's exact test applied where appropriate. Statistical significance: $p < 0.05$ .

These associations should be interpreted in the context of treatment allocation according to clinical need. Patients requiring corticosteroids, supplemental oxygen, or respiratory support generally had greater disease severity than patients who did not require these interventions. Thus, the unadjusted associations between hospital-care interventions and mortality may reflect, at least partly, confounding by indication rather than harmful effects of the treatments themselves.

### Multivariable analysis of demographic and clinical predictors

In the multivariable binary logistic regression model incorporating age group, sex, COPD, and dyspnea, age remained independently associated with hospital survival. Compared with young adults, middle-aged adults had substantially lower adjusted odds of survival (AOR = 0.050; 95% CI: 0.011–0.231; p < 0.001), as did older adults (AOR = 0.030; 95% CI: 0.003–0.278; p = 0.002).

COPD also remained independently associated with outcome (AOR = 6.996; 95% CI: 1.410– 34.697; p = 0.017), although the wide confidence interval indicates considerable imprecision. Dyspnea was independently associated with lower odds of survival (AOR = 0.360; 95% CI: 0.172– 0.754; p = 0.007). In contrast, sex was no longer statistically significant following adjustment (AOR = 0.566; 95% CI: 0.269–1.191; p = 0.134) (Table 5).

**Table 5.** Multivariable binary logistic regression analysis of sociodemographic and clinical factors associated with hospital survival among patients with SARS-CoV-2 infection (N = 348)

| Predictor | Category | B | Adjusted OR | 95% CI | p-value |
| --- | --- | --- | --- | --- | --- |
| Age group | Young adult | Reference | 1.00 | — | — |
|  | Middle adult | -2.990 | 0.050 | 0.011–0.231 | <0.001 |
|  | Old adult | -3.513 | 0.030 | 0.003–0.278 | 0.002 |
| Sex | Female | Reference | 1.00 | — | — |
|  | Male | -0.570 | 0.566 | 0.269–1.191 | 0.134 |
| COPD | No | Reference | 1.00 | — | — |
|  | Yes | 1.945 | 6.996 | 1.410–34.697 | 0.017 |
| Dyspnea | No | Reference | 1.00 | — | — |
|  | Yes | –1.022 | 0.360 | 0.172–0.754 | 0.007 |

*Adjusted ORs were obtained using multivariable binary logistic regression. COPD, chronic obstructive pulmonary disease; CI, confidence interval; OR, odds ratio.*

### Mediation analysis

Three mediation models examined whether patient age group statistically mediated the relationships between steroid therapy, oxygen therapy, respiratory support, and hospital survival.

### Steroid therapy, age group, and survival

Age group significantly mediated the relationship between steroid therapy and survival. After accounting for age group, steroid therapy retained a significant direct association with survival (direct effect, c′ = −2.8670; p = 0.0049). The indirect effect through age group was also statistically significant (a × b = −0.1371; bias-corrected bootstrap 95% CI: −0.3053 to −0.0154). Because the bootstrap confidence interval did not include zero and the direct effect remained statistically significant, the findings were consistent with partial statistical mediation.

Thus, the observed association between steroid therapy and survival comprised both a direct component and an indirect statistical component through differences in patient age group.

### Oxygen therapy, age group, and survival

Oxygen therapy retained a statistically significant direct association with survival after inclusion of age group in the model (c′ = −4.7553; p < 0.001). However, the indirect effect through age group was not statistically significant (a × b = −0.0205; bias-corrected bootstrap 95% CI: −0.1189 to 0.0738). Because the confidence interval included zero, there was no evidence that age group mediated the relationship between oxygen therapy and survival.

### Respiratory support, age group, and survival

No statistically significant indirect effect through age group was identified for the relationship between respiratory support and survival (a × b = 0.0321; bias-corrected bootstrap 95% CI: −0.1537 to 0.2767). Thus, the mediation hypothesis was not supported for respiratory support (Table 6).

**Table 6.** Mediation analysis of the relationships between hospital-care interventions and hospital survival through patient age group (N = 348)

| **Exposure → Mediator → Outcome** | **Direct effect (c')** | **Indirect effect (a×b)** | **Bootstrap 95% CI for indirect effect** | **Direct-effect p-value** | **Mediation** |
| --- | --- | --- | --- | --- | --- |
| Steroid therapy → Age group → Survival | –2.8670 | –0.1371 | –0.3053 to –0.0154 | 0.0049 | **Yes; partial statistical mediation** |
| Oxygen therapy → Age group → Survival | –17.3886* | –0.0478* | –0.2346 to 0.1493* | 0.9828* | No |
| Respiratory support → Age group → Survival | –18.2948* | 0.0321* | –0.1537 to 0.2767* | 0.9764* | No |

Indirect effects estimated using Hayes' PROCESS Macro version 4.2, Model 4, with 5,000 bias-corrected bootstrap resamples. An indirect effect was considered statistically significant when its bootstrap 95% CI excluded zero. Age group should be interpreted as an exploratory statistical mediator, not a causal mediator, because chronological age precedes hospital-care exposure.

### Overall mediation findings

Comparison of the three models showed that the statistical mediating role of age was intervention-specific rather than consistent across all hospital-care interventions. Evidence of mediation was observed only for the steroid therapy–survival relationship, for which age group demonstrated a significant indirect effect alongside a significant direct effect. No evidence of mediation was observed for oxygen therapy or respiratory support (Fig 1).

**Fig 1.**
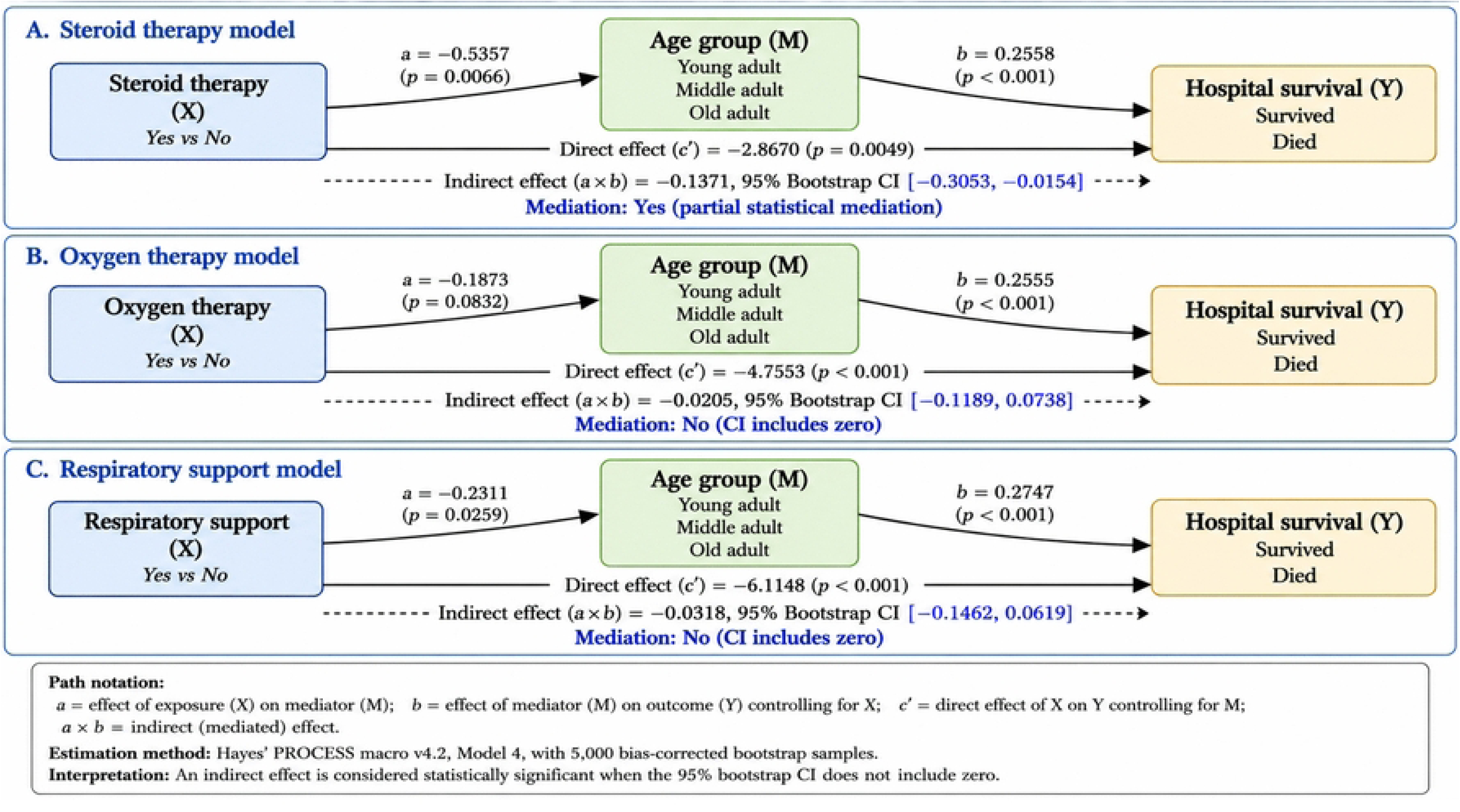
Statistical mediation models examining the mediating role of patient age group in the associations between hospital-care interventions and hospital survival

Given the observational design and the fact that chronological age necessarily precedes the hospital-care interventions, these findings represent statistical decomposition of the observed associations and should not be interpreted as evidence that the interventions causally operated through or altered patient age.

## 4. DISCUSSION

This prospective observational study provides context-specific evidence on the clinico-epidemiological characteristics, hospital-care interventions, and survival outcomes of adults hospitalized with SARS-CoV-2 infection in a tertiary care hospital in Nepal. Approximately four-fifths of the participants survived to hospital discharge, while one-fifth died. Older age groups constituted a substantial proportion of hospitalized patients, hypertension and diabetes were the most prevalent comorbidities, and fever, cough, and dyspnea were the dominant presenting symptoms. Age, sex, COPD, dyspnea, and major hospital-care interventions showed unadjusted associations with survival, while age, COPD, and dyspnea remained independently associated with outcome in the demographic and clinical multivariable model. Most importantly, the mediation analyses demonstrated an intervention-specific pattern: patient age group statistically mediated the association between steroid therapy and survival, whereas no significant indirect effect through age was identified for oxygen therapy or respiratory support. These findings highlight the combined importance of baseline vulnerability, clinical presentation, treatment requirements, and age-related heterogeneity in understanding hospital outcomes of COVID-19.

### Clinico-epidemiological characteristics and hospital survival

The predominance of middle-aged and older adults in the study is consistent with the extensive evidence demonstrating a strong age gradient in severe COVID-19, hospitalization, and mortality. The OpenSAFELY study involving more than 17 million adults in England identified increasing age as one of the strongest factors associated with COVID-19-related death [3]. Similar findings were reported by Zhou et al. among hospitalized adults in Wuhan, where older age independently increased the odds of in-hospital death [14]. Age-related vulnerability to severe COVID-19 is biologically plausible and probably reflects multiple interacting mechanisms, including immunosenescence, chronic low-grade inflammation, reduced physiological reserve, endothelial dysfunction, and the increasing burden of chronic diseases with advancing age [15,16].

The slight male predominance observed in the present study is also consistent with several large COVID-19 cohorts. OpenSAFELY identified male sex as independently associated with COVID- 19-related mortality [3], while studies of critically ill patients have similarly reported a predominance of men among patients requiring intensive care [17]. Several mechanisms have been proposed, including sex-related differences in immune response, hormonal regulation, cardiometabolic risk, smoking exposure, and health-seeking behaviour. In the present study, however, sex did not retain statistical significance after multivariable adjustment. This suggests that the unadjusted association between sex and survival may have been partly explained by differences in age, comorbidity, clinical presentation, or other characteristics rather than an independent effect of sex itself.

Hypertension and diabetes mellitus were the most frequent comorbidities in this cohort. This pattern is broadly consistent with hospital-based studies from Nepal and elsewhere [8,9,14]. A multicentre assessment of COVID-19 mortality in Nepal similarly identified hypertension and diabetes among the most common underlying conditions among patients who died [8].

Nevertheless, neither hypertension nor diabetes showed an independent association with survival in the present demographic and clinical model. This finding should not be interpreted as evidence that these conditions are unimportant in COVID-19. Their effects may be partly mediated or confounded by age, multimorbidity, disease severity, obesity, cardiovascular disease, treatment, and other factors. Large population studies have demonstrated associations between several cardiometabolic conditions and COVID-19 mortality, although the magnitude of these associations changes substantially following adjustment for age and other covariates [3].

Fever and cough were the most common presenting symptoms, while dyspnea was present in nearly half of the participants. These findings are compatible with the established clinical spectrum of symptomatic COVID-19. Of particular importance, dyspnea was strongly associated with mortality and remained independently associated with lower odds of survival after adjustment. Dyspnea is a clinically meaningful marker because it may reflect pulmonary involvement, hypoxemia, impaired gas exchange, or progression toward respiratory failure. In a Nepalese hospital mortality review, shortness of breath was reported in 92% of patients who died [9]. The independent association observed in the present study therefore reinforces the importance of respiratory symptoms as readily identifiable indicators for early clinical risk assessment, particularly where sophisticated prognostic tools may not be immediately available.

### Age as an independent determinant of survival

Age remained one of the strongest independent demographic predictors of hospital survival. Compared with young adults, both middle-aged and older adults demonstrated markedly lower adjusted odds of survival. This finding is consistent with international evidence demonstrating that advancing age is a major determinant of COVID-19 mortality [3,14,17].

Several mechanisms may account for the strong relationship between age and adverse outcome. Ageing is associated with progressive impairment of innate and adaptive immunity, diminished T-cell function, dysregulated inflammatory responses, endothelial dysfunction, and reduced pulmonary and cardiovascular reserve [15,16]. Older adults are also more likely to have multimorbidity and frailty, which may limit their ability to tolerate severe infection and prolonged hospitalization. The effect of chronological age therefore probably represents a combination of biological ageing, accumulated disease burden, functional reserve, and differences in response to acute physiological stress rather than age alone.

From a public health perspective, the strong age gradient has implications beyond individual prognosis. During periods of high hospital demand, age—together with clinical severity and comorbidity—can contribute to early identification of populations requiring closer observation and timely escalation of care. However, age should not be used in isolation to restrict access to treatment. Risk stratification should remain multidimensional and should incorporate clinical status, comorbidities, physiological parameters, patient preferences, and expected treatment benefit.

### COPD and survival outcome

COPD remained significantly associated with survival after adjustment, although the estimate was imprecise, as reflected by the wide confidence interval. The relationship between chronic respiratory disease and adverse COVID-19 outcomes is biologically plausible because patients with COPD commonly have impaired pulmonary reserve, chronic airway inflammation, altered immune responses, and reduced ability to compensate for an acute respiratory infection.

International evidence generally identifies COPD as an important risk factor for adverse COVID- 19 outcomes. A systematic review and meta-analysis of 59 studies reported that COPD was associated with increased odds of hospitalization, intensive care admission, and mortality among patients with COVID-19 [18]. This reinforces the importance of recognizing chronic respiratory disease during admission assessment.

Nevertheless, the direction of the adjusted COPD estimate in the present dataset should be interpreted cautiously. The observed AOR was greater than one for the coded survival outcome, whereas the crude data showed an unusual distribution of deaths among participants with and without documented COPD. This may reflect outcome coding, small cell counts, selection effects, documentation patterns, or characteristics of the fitted model. Consequently, although COPD emerged as statistically significant, its effect size and direction should be verified against the original coding and analytical dataset before the manuscript is submitted.

### Hospital-care interventions and survival

Steroid therapy, supplemental oxygen, and respiratory support were each strongly associated with survival in the unadjusted analyses. Mortality was greater among patients receiving these interventions than among those who did not. These findings should not be interpreted as indicating that the interventions increased mortality. In an observational hospital cohort, treatments are administered according to clinical need rather than randomly allocated. Patients receiving oxygen, corticosteroids, or advanced respiratory support are therefore generally more severely ill than untreated patients. This creates a classic problem of confounding by indication, in which the severity that prompted treatment is itself strongly related to the outcome.

This distinction is particularly important for corticosteroids. The RECOVERY randomized trial demonstrated that dexamethasone reduced 28-day mortality among hospitalized patients receiving invasive mechanical ventilation or supplemental oxygen but did not provide benefit among patients who did not require respiratory support [5]. The apparently poorer crude survival among steroid-treated participants in the present study therefore should not be interpreted as contradicting the RECOVERY trial. Rather, it likely reflects differences in underlying disease severity, treatment indications, timing, dose, duration, and other clinical characteristics that could not be fully accounted for in the observational analysis.

A similar interpretation applies to oxygen therapy and respiratory support. Supplemental oxygen is a treatment for hypoxemia, while non-invasive or invasive respiratory support is generally initiated when respiratory compromise becomes more severe. Consequently, the requirement for these interventions functions partly as a marker of illness severity. The finding that no deaths occurred among patients who did not require oxygen further illustrates this issue: patients who maintained adequate oxygenation were inherently at substantially lower clinical risk than those who developed an oxygen requirement.

These findings have an important health-system dimension. Access to reliable medical oxygen, respiratory support, appropriately trained personnel, monitoring equipment, and critical care capacity became a central health-system challenge during the pandemic, particularly in LMICs. Ensuring timely and equitable access to these resources remains relevant for preparedness for future respiratory infectious disease emergencies.

### Mediating role of age in the steroid therapy–survival relationship

The principal analytical contribution of this study was the examination of whether age statistically mediated the relationships between selected hospital-care interventions and survival. In the steroid therapy model, both the direct effect of steroid therapy and the indirect effect through patient age group were statistically significant. The bootstrap confidence interval for the indirect effect excluded zero, supporting partial statistical mediation.

This finding indicates that the observed steroid therapy–survival association was statistically decomposed into a component that remained after accounting for age and an additional component associated with differences across age groups. In other words, age explained part, but not all, of the statistical relationship between steroid exposure and survival. The finding is potentially important because corticosteroid treatment is closely linked to clinical severity and oxygen requirement, while age is strongly associated with both disease severity and mortality.

The result should, however, be interpreted with substantial methodological caution. A conventional causal mediator must generally occur after the exposure and before the outcome. Chronological age exists before hospitalization and necessarily precedes administration of corticosteroids. Steroid therapy therefore cannot cause a patient’s age. Accordingly, the significant indirect effect observed in this study does not demonstrate that steroid therapy affects survival by changing age, nor does it establish a biological causal mediation mechanism.

Rather, age in this model should be understood as an exploratory statistical mediator or pathway variable that captures age-related heterogeneity in the observed steroid–survival relationship. This distinction is essential. Hayes’ regression-based mediation framework provides methods for estimating direct and indirect statistical effects [10], but the causal interpretation of those effects depends on appropriate temporal ordering and substantive assumptions about confounding. Modern causal mediation literature similarly emphasizes that statistical evidence of an indirect effect alone is insufficient to establish a causal mechanism [19,20].

The partial statistical mediation nevertheless raises an important clinical question: why might the observed relationship between corticosteroid exposure and outcome vary according to age? Older patients may differ from younger patients in baseline immune function, inflammatory response, comorbidity burden, physiological reserve, disease severity, treatment thresholds, and tolerance of complications [15,16]. Corticosteroid use may also be more frequent among patients with severe inflammatory pulmonary disease, creating complex interactions among age, indication for treatment, severity, and survival. These mechanisms cannot be distinguished by the present mediation model but provide hypotheses for future investigation.

The RECOVERY trial further demonstrates why treatment context is important: corticosteroid benefit differed according to respiratory support at randomization [5]. Therefore, future studies examining age-related heterogeneity in corticosteroid effectiveness should preferably evaluate age as a potential effect modifier, stratification factor, or confounder, alongside formal causal mediation models using temporally appropriate mediators such as inflammatory response, oxygen requirement, disease progression, or organ dysfunction.

### Absence of age mediation for oxygen therapy and respiratory support

In contrast to steroid therapy, the indirect effects of oxygen therapy and respiratory support through age were not statistically significant. The bootstrap confidence intervals included zero in both models, providing no evidence that age statistically mediated these intervention–survival relationships.

This finding is informative because it demonstrates that age did not function as a universal intermediary across all treatment variables. The need for oxygen and respiratory support is determined predominantly by immediate physiological deterioration, particularly hypoxemia and respiratory failure. Although older age increases the overall risk of severe disease, the pathway linking respiratory support requirements to mortality may be more directly determined by acute pulmonary dysfunction, disease severity, organ failure, treatment timing, and critical care complications than by age itself.

The contrast among the three mediation models also illustrates an important methodological principle: a variable should not be assumed to mediate every exposure–outcome association merely because it is strongly associated with the outcome. Mediation is pathway-specific. The significant indirect effect observed for steroid therapy but not for oxygen or respiratory support suggests heterogeneity in the statistical relationships among treatment, age, and survival.

However, the non-significant indirect effects should not be interpreted as evidence that age is clinically irrelevant to oxygen or respiratory-support outcomes. Age remains a major prognostic factor for COVID-19 mortality. Rather, within the statistical specification and sample of the present study, there was insufficient evidence that the associations of oxygen therapy or respiratory support with survival operated indirectly through the categorized age variable.

### Interpretation of mediation findings in an observational study

The mediation results warrant careful distinction between statistical mediation and causal mediation. Mediation analysis is frequently used to decompose an exposure–outcome association into direct and indirect components, and bootstrap estimation is advantageous because it does not require the sampling distribution of the indirect effect to be normal [10,20]. Nevertheless, causal interpretation requires additional assumptions, including appropriate temporal sequencing and adequate control of exposure–mediator, mediator–outcome, and exposure–outcome confounding [19].

These assumptions are difficult to satisfy in the present observational setting. Age preceded all hospital-care interventions, disease severity influenced treatment allocation, and important factors such as frailty, vaccination status, timing of treatment, oxygen saturation, detailed severity indices, corticosteroid dose and duration, and ventilatory parameters were not incorporated into the mediation models. Consequently, the mediation results should be viewed primarily as exploratory statistical decomposition rather than evidence of a causal pathway.

This cautious interpretation does not diminish the value of the analysis. Rather, it defines precisely what can and cannot be inferred from it. The finding that age accounted statistically for part of the steroid–survival association generates a hypothesis regarding age-related treatment heterogeneity that can be evaluated in future prospective multicentre studies using temporally ordered variables and causal mediation methods.

### Public health and health-system implications

The findings have several implications for public health practice in Nepal and other resource-constrained settings. First, age and readily identifiable clinical characteristics such as dyspnea can support early recognition of patients at increased risk of deterioration. Risk assessment based on information available at admission is particularly useful in settings where advanced diagnostic resources may be constrained.

Second, the observed relationships between oxygen therapy, respiratory support, and mortality underscore the importance of interpreting treatment utilization as an indicator of both healthcare need and disease severity. During major respiratory outbreaks, health systems require reliable oxygen supply chains, functional delivery systems, respiratory-support equipment, appropriately trained healthcare workers, and escalation pathways linking general wards with critical care. Strengthening these components is relevant not only for COVID-19 but also for future epidemics of severe respiratory infection.

Third, the findings support an age-sensitive rather than age-exclusive approach to clinical and public health decision-making. Older adults constitute a high-risk population, but chronological age should be considered alongside disease severity, comorbidities, physiological status, and treatment requirements. Such multidimensional assessment can support equitable prioritization of monitoring and resources without reducing clinical decisions to age alone.

Finally, the study demonstrates the value of locally generated evidence. Patterns of hospital presentation, treatment availability, referral, critical care capacity, and population vulnerability differ between countries and health systems. Evidence from Nepal is therefore important for complementing large studies from high-income settings and informing preparedness strategies appropriate to the national context.

### Strengths and limitations

This study has several strengths. Its prospective observational design allowed systematic collection of information during hospitalization rather than relying exclusively on retrospective records. Consecutive recruitment reduced discretionary selection of eligible participants, and all 348 participants were followed to a definitive hospital outcome, eliminating loss to follow-up for the primary outcome. The analysis combined descriptive epidemiology, unadjusted comparisons, multivariable regression, and mediation analysis, providing complementary perspectives on the relationships between patient characteristics, hospital care, and survival. The use of bootstrap confidence intervals for the indirect effects also strengthened statistical estimation of the mediation models.

Several limitations should nevertheless be considered. First, this was a single-centre hospital-based study; therefore, the findings may not be directly generalizable to community cases, primary or secondary hospitals, other regions of Nepal, or healthcare systems with substantially different resources and referral patterns. Second, the observational design prevents causal conclusions regarding treatment effects. Steroid therapy, oxygen therapy, and respiratory support were prescribed according to clinical indication, making confounding by indication particularly important.

Third, residual confounding is likely. Potentially relevant factors such as vaccination status, viral variant, frailty, body mass index, socioeconomic status, timing from symptom onset to admission, baseline oxygen saturation, detailed disease-severity measures, corticosteroid type/dose/timing, and detailed ventilatory parameters were unavailable or were not incorporated into the present analysis. Fourth, routinely documented hospital data may be subject to measurement and classification error.

Fifth, categorizing age may have reduced information and statistical power compared with modelling chronological age continuously and may have introduced dependence on the selected category thresholds. Sixth, the use of age as a mediator has a fundamental temporal limitation because age necessarily precedes the hospital-care exposures. The mediation results therefore represent statistical indirect effects rather than confirmed causal mediation. Future studies should examine age as a confounder or effect modifier and use temporally plausible post-exposure variables when testing causal mediation.

Seventh, some analyses involved small cell counts and wide confidence intervals, particularly for COPD, which limits precision. The coding and direction of the COPD estimate should be verified before final submission. Finally, the outcome was restricted to survival at hospital discharge. Long-term mortality, readmission, functional status, and post-COVID outcomes were beyond the scope of the study.

Despite these limitations, the study adds prospective evidence from Nepal regarding the relationship between patient vulnerability, clinical presentation, hospital-care requirements, and short-term survival and provides an exploratory assessment of age-related statistical pathways in treatment–outcome relationships.

### Recommendations

Based on the findings of this study, the following recommendations are proposed:

1. Strengthen age-sensitive clinical risk assessment: Age, together with presenting clinical characteristics, particularly dyspnea, should be considered in the early risk assessment of patients hospitalized with SARS-CoV-2 infection. Patients with greater age-related vulnerability or respiratory compromise may require closer monitoring and timely escalation of supportive care.
2. Strengthen respiratory-care and health-system preparedness: Healthcare institutions, particularly in resource-constrained settings, should ensure reliable medical oxygen systems, adequate respiratory-support and critical-care capacity, appropriate monitoring facilities, trained healthcare personnel, and clearly defined pathways for escalation of care. Such preparedness may also strengthen responses to future severe respiratory infectious disease outbreaks.
3. Interpret hospital-care intervention findings cautiously: The observed associations of steroid therapy, oxygen therapy, and respiratory support with hospital survival should not be interpreted as evidence of beneficial or harmful treatment effects because these interventions were provided according to clinical need and were likely influenced by disease severity and confounding by indication. Future studies should account for disease severity, treatment indication, timing, dose and duration of therapy, and other relevant confounders.
4. Further investigate the role of age in treatment–outcome relationships: The significant statistical mediation of the steroid therapy–survival relationship through age group warrants further investigation. However, because chronological age precedes treatment exposure, this finding should not be interpreted as a causal mediating pathway. Future studies should examine age as a potential effect modifier, stratification factor, or confounder and assess whether treatment–outcome relationships differ across age groups.
5. Use temporally appropriate mediation and advanced analytical approaches: Future mediation studies should prioritize clinically plausible post-exposure mediators, such as changes in disease severity, oxygen requirement, organ dysfunction, or treatment response. Interaction analysis, appropriately designed causal mediation analysis, and other advanced modelling approaches may provide a more robust understanding of the pathways linking hospital-care interventions with clinical outcomes.
6. Conduct larger multicentre studies with broader outcomes: Larger prospective studies across different healthcare settings in Nepal and other low- and middle-income countries are recommended to validate these findings and improve generalizability. Future studies should consider age as both a continuous and categorical variable and extend outcome assessment beyond hospital survival to include hospital and intensive care length of stay, readmission, functional recovery, post-COVID morbidity, quality of life, and longer-term mortality.

## CONCLUSION

Among adults hospitalized with SARS-CoV-2 infection in a tertiary care hospital in Nepal, mortality remained substantial, with approximately one in five patients dying before hospital discharge. Age and dyspnea emerged as important demographic and clinical indicators of adverse outcome, while COPD demonstrated an adjusted association that requires cautious interpretation and verification because of imprecision and coding considerations. Hospital-care interventions were strongly associated with outcome in unadjusted analyses, but these associations should not be interpreted as treatment effects because of confounding by indication.

Age group statistically mediated the association between steroid therapy and survival but did not mediate the associations involving oxygen therapy or respiratory support. This intervention-specific finding suggests age-related heterogeneity in the observed steroid–survival relationship; however, because age precedes treatment, the indirect effect represents exploratory statistical rather than causal mediation.

These findings support early identification of high-risk hospitalized patients, age-sensitive clinical assessment, and continued strengthening of oxygen, respiratory-support, and critical-care capacity in resource-constrained health systems. Future multicentre prospective studies should incorporate detailed measures of disease severity and treatment timing and apply temporally appropriate causal mediation or effect-modification analyses to clarify how age influences treatment–outcome relationships.

## Data Availability

All data are in the manuscript and/or supporting information files.

## ACKNOWLEDGMENTS

The authors would like to express their sincere gratitude to **Prof. Dr. Harish Chandra Neupane** and **Prof. Dr. Gopendra Prasad Deo** for their invaluable support and facilitation during the data collection process. We also extend our sincere appreciation to the in-charges and healthcare staff of the COVID Intensive Care Unit and COVID Ward for their cooperation and support throughout the study. Finally, we are deeply grateful to all the patients who participated in this study and generously contributed their time and information, without whom this research would not have been possible.

## SUPPORTING INFORMATION LEGENDS

S1 Fig 1. Statistical mediation models examining the mediating role of patient age group in the associations between hospital-care interventions and hospital survival.

S1 Data. Study dataset used for the statistical analyses of the Clinico-Epidemiological Characteristics, Hospital Care Interventions, and Clinical Outcomes of Patients with SARS-CoV- 2 in Nepal: The Mediating Role of Age. (PDF)

S1 Text. Global inclusivity questionnaire completed in accordance with PLOS Global Public Health reporting requirements. (DOCX)

## SUBMISSION DECLARATIONS

### Data Availability Statement

All data are in the manuscript and/or supporting information files.

## Abbreviations

AOR: Adjusted Odds Ratio
CI: Confidence Interval
CMCTH: Chitwan Medical College Teaching Hospital
COPD: Chronic Obstructive Pulmonary Disease
COVID-19: Coronavirus Disease 2019
ICU: Intensive Care Unit
LMICs: Low- and Middle-Income Countries
NHRC: Nepal Health Research Council
OR: Odds Ratio
RT-PCR: Reverse Transcription Polymerase Chain Reaction
SARS-CoV-2: Severe Acute Respiratory Syndrome Coronavirus 2
SPSS: Statistical Package for the Social Sciences
STROBE: Strengthening the Reporting of Observational Studies in Epidemiology
WHO: World Health Organization.

## Author Contributions Mamata Sharma Neupane

**Roles** Conceptualization, Data curation, Formal analysis, Funding acquisition, Investigation, Methodology, Project administration, Resources, Software, Supervision, Validation, Visualization, Writing – original draft, Writing – review & editing

**Hafizah Che Hassan**

**Roles** Conceptualization, Data curation, Formal analysis, Methodology, Supervision, Validation, Writing – review & editing

**Surendra Uranw**

**Roles** Conceptualization, Data curation, Formal analysis, Methodology, Software, Supervision, Validation, Writing – review & editing

**Raj Kumar Mehta**

**Roles** Conceptualization, Data curation, Formal analysis, Methodology, Resources, Software, Validation, Writing – review & editing

